# Trends in Pedestrian-related mortality in the United States; 1999-2020, A CDC-WONDER analysis

**DOI:** 10.1101/2025.07.16.25331672

**Authors:** Syed Usama Ashraf, Anoud Khan, Aryan Tareen, Muntaha Irfan, Arooba Hameed Shaikh, Syeda Mashal Ushna Danish, Ayesha Imran, Amina Mujahid

## Abstract

**Objective:** Pedestrian mortality in the United States has increased seven times faster than the population growth from 2019 to 2023, according to a Governors Highway Safety Association report. This alarming trend highlights the need to study pedestrian mortality patterns, stratified by gender, race/ethnicity, age group, and state-specific characteristics, along with an exploration of contributing factors driving this surge.

**Methods:** The Centers for Disease Control and Prevention Wide-Ranging Online Data for Epidemiologic Research (CDC-WONDER) database was used to extract pedestrian mortality data from 1999 to 2020. Age-Adjusted Mortality Rates (AAMRs) per 100,000 population and Annual Percentage Changes (APCs) with 95% confidence intervals (CIs) were calculated. Joinpoint regression analysis was employed to assess the trends across various demographic and regional subgroups.

**Results:** A total of 140,280 pedestrian deaths occurred in the US between 1999 and 2020. The overall AAMR increased from 2.21 in 1999 to 2.32 in 2020. A steep rise in the APC (3.11) was observed from 2009 to 2020. Men consistently had higher AAMRs than women, while non-Hispanic (NH) American Indians or Alaska Natives had the highest AAMR among races. Individuals aged 35–44 years exhibited the highest APC (6.92) between 2011 and 2020. States in the 90th percentile (Arizona, Florida, New Mexico) had triple AAMRs compared to those in the 10th percentile. Rural areas had the highest APC (3.23) from 2011-2020.

**Conclusion:** Pedestrian mortality rates in the United States have been rising for over a decade. Enhanced public safety interventions and efforts to address disparities based on race, age, gender, and geographic location are essential to curb the growing burden of pedestrian deaths.

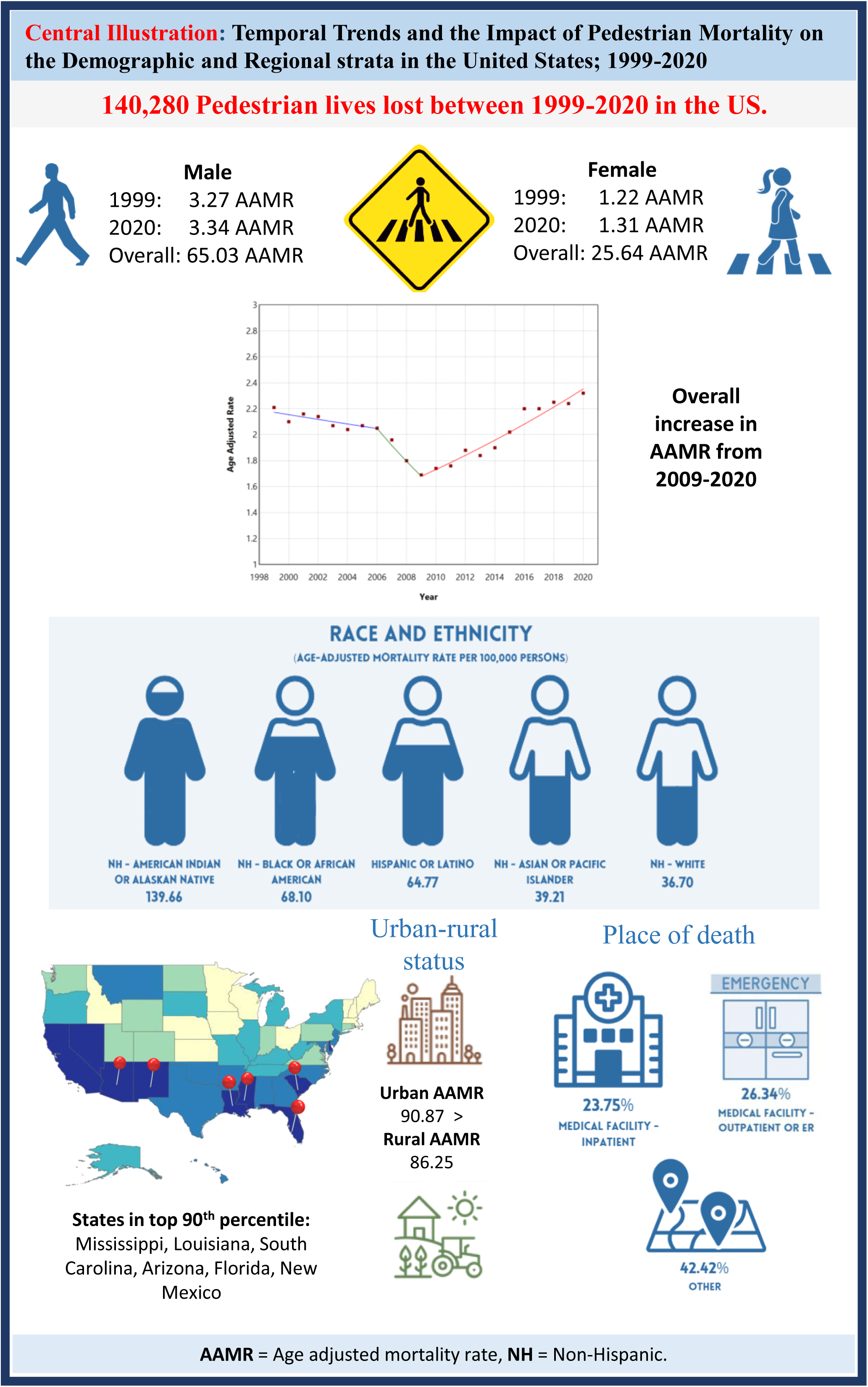

## INTRODUCTION

Pedestrian mortality has been silently rising each year, yet it remains one of the most overlooked causes of death in the United States. Globally, pedestrians account for a third of the 1.2 million traffic fatalities each year, and the challenges continue to escalate. [1] According to the World Health Organization, the global death toll from traffic injuries surpasses 1.35 million annually, with pedestrians and cyclists accounting for 26% of these deaths. [2] In the United States alone, an estimated 80,000 to 120,000 pedestrians are injured and 4,600 to 4,900 die annually from motor vehicle crashes, making them the most vulnerable among all road users. [3]

Pedestrian mortality in the United States reached a 40-year high in 2021, nearing 8,000 deaths; equating to one death every 66 minutes. [4,5] The total medical costs associated with pedestrian mortality are estimated at $9.71 million in 2022. [6] This growing trend is more than a collection of statistics; it reflects deeper structural issues, including changes in urban landscapes and transportation patterns. Increased urban density and a surge in heavier vehicles such as SUVs and trucks are more likely to result in fatal pedestrian crashes. [3] Among all racial and ethnic groups, pedestrian mortality remains significantly higher for American Indians and Alaska Natives. Of those aged 1–44 years; pedestrian injuries are the third leading cause of death. [7] Contributing factors include alcohol use, poverty, and limited access to education. [7]

Simultaneously, cultural shifts toward more active lifestyles and environmentally conscious transportation have increased pedestrian activity, thereby testing traditional assumptions about road safety. [8] Alarmingly, a disproportionate number of pedestrian deaths occur in low-light conditions, highlighting the urgent need to examine urban lighting and visibility standards. [9] Physical activity is well-documented to reduce the risk of chronic diseases and early mortality while promoting overall well-being. However, the rising pedestrian death toll paradoxically threatens these public health benefits, as growing safety concerns may discourage individuals from walking. [10]

At present, there is a dearth of literature concerning trends in pedestrian-related mortality, which warrants extensive analysis. In this 21-year study, our study aims to map the undulating patterns of pedestrian mortality across two-decades (1999-2020). By studying demographic and regional trends we also aim to provide insights that outline evidence-based interventions to reduce pedestrian fatalities and foster safer streets.

## METHODS

### Study Setting and Population

A retrospective analysis was performed to study the pedestrian mortality trends in the United States. The total duration of the study comprises of data collected from 1999 to 2020, that were obtained from Centers for Disease Control and Prevention Wide-Ranging Online Data for Epidemiologic Research (CDC-WONDER) database. The Multiple Cause-of-Death public use records were reviewed to select pedestrian deaths, which identified pedestrian wounded in traffic injury as a contributory or underlying cause of death. The analysis utilized codes from the International Statistical Classification of Diseases and Related Health Problems, 10th Revision (ICD-10), specifically V01-V09 (Pedestrians injured in transport crashes). The same ICD codes have been previously used to recognize pedestrian mortality in administrative databases. [11] These codes classify various types of pedestrian injuries: V01 for pedestrians injured in collisions with pedal cycles, V02 for pedestrians injured in collisions with two- or three-wheeled motor vehicles, V03 for pedestrians injured in collisions with cars, pick-up trucks, or vans, V04 for pedestrians injured in collisions with heavy transport vehicles or buses, V05 for pedestrians injured in collisions with railway trains or vehicles, V06 for pedestrians injured in collisions with other non-motor vehicles, and V09 for pedestrians involved in non-traffic injuries with other and unspecified motor vehicles. This study was exempt from local institutional review board approval as it utilized de-identified, publicly available government-issued data. The study adhered to the STROBE (Strengthening the Reporting of Observational Studies in Epidemiology) guidelines for reporting.

### Data Abstraction

#### Inclusion criteria

The aim of this study was to analyse mortality trends among individuals of all ages. Therefore, data was abstracted and analysed based on overall AAMR, gender, race/ethnicity, 10-year age-group, states, urban-rural status and place of death variables. Genders included were male and female as per the death certificates. Race/ethnicity was classified as non-Hispanic (NH) White, NH-Black or African American, Hispanic or Latino, NH-American Indian or Alaska Native, and NH-Asian or Pacific Islander. 10-year age groups were classified as 25-34, 35-44, 45-54, 55-64, 65-74, 75-84, and 85+ years. Place of death variables for medical facilities were categorized into medical facility (outpatient, emergency room, inpatient, death on arrival, or status unknown), home, hospice, nursing home/ long-term care facility, ‘other’ and ‘unknown’ locations. This analysis is based on death certificate data which has also been used in earlier CDC-WONDER database analyses. [12] The National Center for Health Statistics Urban-Rural Classification Scheme, based on the 2013 U.S. Census, was used to categorize the population into urban and rural areas. Urban areas included large metropolitan regions with populations of 1 million or more, as well as medium/small metropolitan areas with populations ranging from 50,000 to 999,999. Rural areas were defined as those with populations under 50,000. [13]

#### Exclusion criteria

1059 deaths were not included in the final data when stratifying for race/ethnicity as it was reported in the ‘not stated’ category of race/ethnicity. Also mortalities occurring in 0-24 year of age when stratifying for 10-year age groups were also excluded.

### Statistical Analysis

To explore nation wide trends in pedestrian mortality from 1999 to 2020, we calculated crude-mortality rates (CMRs) and age-adjusted mortality rates (AAMRs) per 100,000 population with 95% CI, stratified for demographic and geographic variables. CMRs were derived by dividing the number of pedestrian deaths by the corresponding year’s U.S. population. AAMRs were standardized to the year 2000 U.S. population for consistency. [14] To analyze annual trends, we utilized the Joinpoint Regression Program (version 5.0, National Cancer Institute) to determine the annual percent change (APC) in AAMR with 95% CIs. [15, **Table A1**] This method identifies significant changes in mortality rates over time by fitting log-linear regression models to periods of temporal variation. Changes in the Annual Percentage Change (APC) were classified as either increasing or decreasing if the slope was significantly different from zero, as determined by two-tailed t-tests. A p-value of less than 0.05 was considered indicative of statistical significance.

## RESULTS

A total of 140,280 pedestrian fatalities were reported between 1999-2020 in the United States **(Table A2).** Data was divided into three categories: demographic (gender, race/ethnicity, 10-year age groups), geographic (states and urban-rural status) and place of death variables. Data on place of death was available for 140,280 deaths. Of which, 26.34% occurring in outpatient or ER, 23.75% inpatient, dead on arrival 3.56%, 2.01% at home, 0.93% in nursing and long-term care, 0.50% in hospice and 0.41% in unknown locations. The highest percentage of 42.42% was seen in ‘other’ locations. **(Table A3)**

### Annual Trends for Pedestrian-related AAMR

The overall trend of AAMR has steadily decreased from 1999 to 2006 (APC: −0.84; 95% CI −1.96 to 0.28) which was then followed by a sharp decline from 2006 to 2009 (APC: −6.39; 95% CI −13.56 to 1.38). An alarming increase has been observed from 2009-2020 (APC: 3.11; 95% CI 2.60 to 3.62) **(Figure 1 and Table A1)**. The overall AAMR for Pedestrian-related deaths were 2.21 in 1999 and 2.32 in 2020. **(Table A4).**

**Figure 1:**
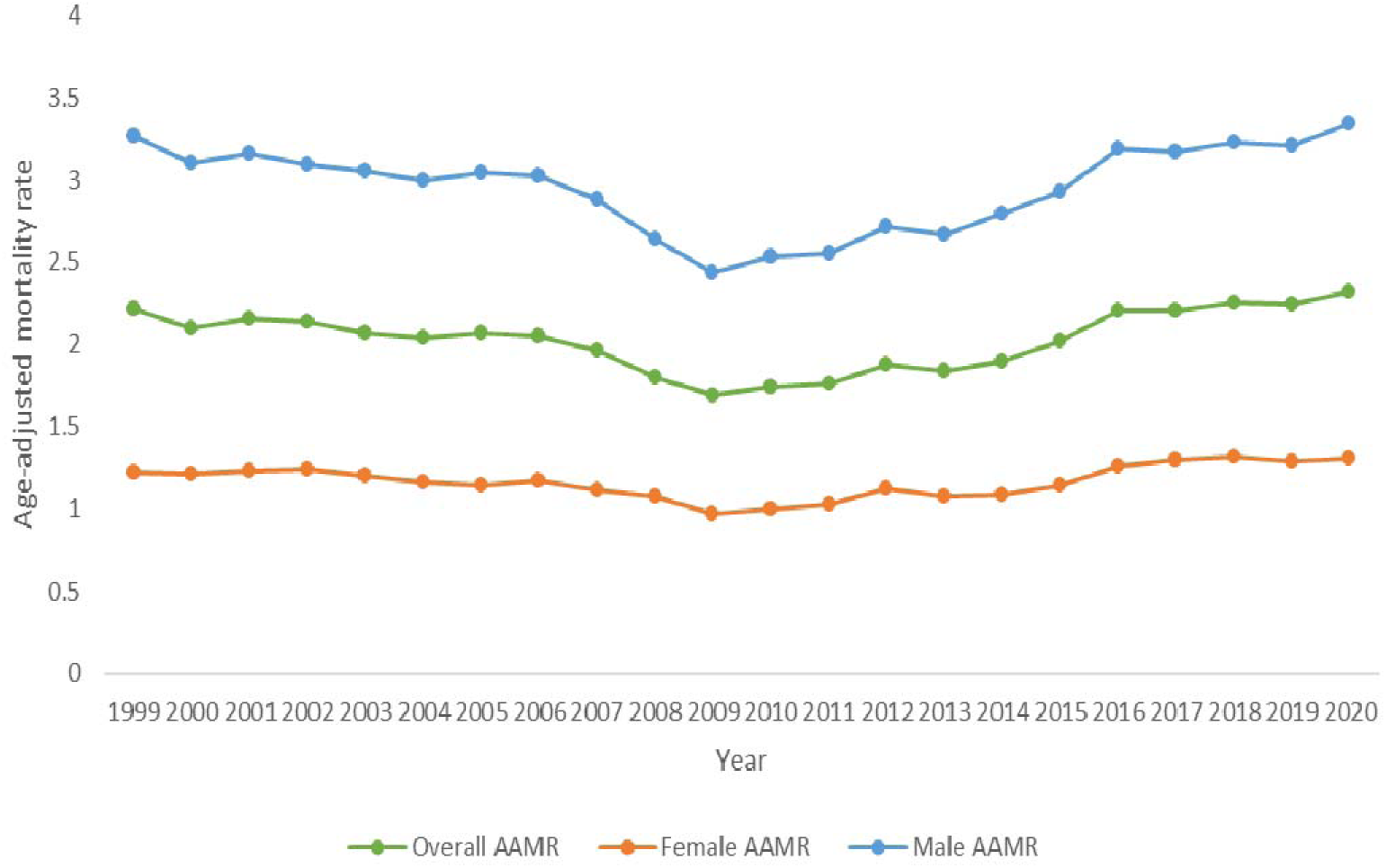
Pedestrian overall and gender stratified age-adjusted mortality rate in the United States; 1999-2020

### Pedestrian-related AAMR stratified by gender

It was observed that males had a significantly higher mortality compared with females, with a total of 98,584 deaths in males and 41,696 deaths in females. **(Table A2)**. In males, the AAMR was 3.27 in 1999 and 3.34 in 2020 **(Table A4).** From 1999 to 2006, the Age-Adjusted Mortality Rate (AAMR) in men initially exhibited a steady decline, with an Annual Percentage Change (APC) of −0.82 (95% CI: −1.57 to 0.07). This was followed by a sharp decrease in AAMR until 2009 (APC: −6.33; 95% CI: −11.03 to −1.38). Subsequently, the trend showed a modest increase until 2013 (APC: 2.0; 95% CI: −0.68 to 4.83), followed by a more pronounced rise until 2016 (APC: 5.36; 95% CI: 0.27 to 10.72). The rate then continued to increase at a slower pace through 2020 (APC: 1.23; 95% CI: −0.30 to 2.78). **(Figure 1 and Table A1)**

In females the AAMR was 1.22 in 1999 and 1.31 in 2020. **(Table A4)**. A decreasing pattern is observed from 1999 to 2006 with an associated APC of −0.84 (95% CI −1.96 to 0.29), followed by a sharp decline (APC −5.28, 95% CI −13.62 to 3.86) till 2009. It is then marked by an increasing trend till 2014 with an APC of 2.20 (95% CI −0.88 to 5.37), followed by a significantly increasing trend till 2017 with an APC of 5.93 (95% CI −2.60 to 15.21) and till 2020 a decreasing trend was observed with an APC of −0.16 (95% CI −3.90 to 3.72) **(Figure 1 and Table A1)**

### Pedestrian-related AAMR stratified by race/ethnicity

Our results demonstrate the absolute number of deaths were highest in NH-Whites (78,021) followed by NH-Black or African American (26,288), Hispanics or Latino (26,124), NH-Asian or Pacific Islander (5336) and NH-American Indian or Alaska Native (3452). **(Table A2)** Data when stratified by race/ethnicity, demonstrated the highest AAMR in NH-American Indian or Alaska Native (139.66), followed by NH-Black or African American (68.10), Hispanic or Latino (64.77), NH- Asian or Pacific Islander (39.21) and NH- White (36.70). **(Table A5)**

NH-American Indians or Alaskan native had an AAMR of 7.45 in 1999 and 6.78 in 2020. The trend in mortality was seen to decrease from 1999 to 2011 (APC; −1.65; 95% CI −3.02 to −0.26) which was then followed by a progressive increase till 2020 (APC; 2.79 95% CI 0.80 to 4.82). **(Table A1, Figure 2, Table A5)**

**Figure 2:**
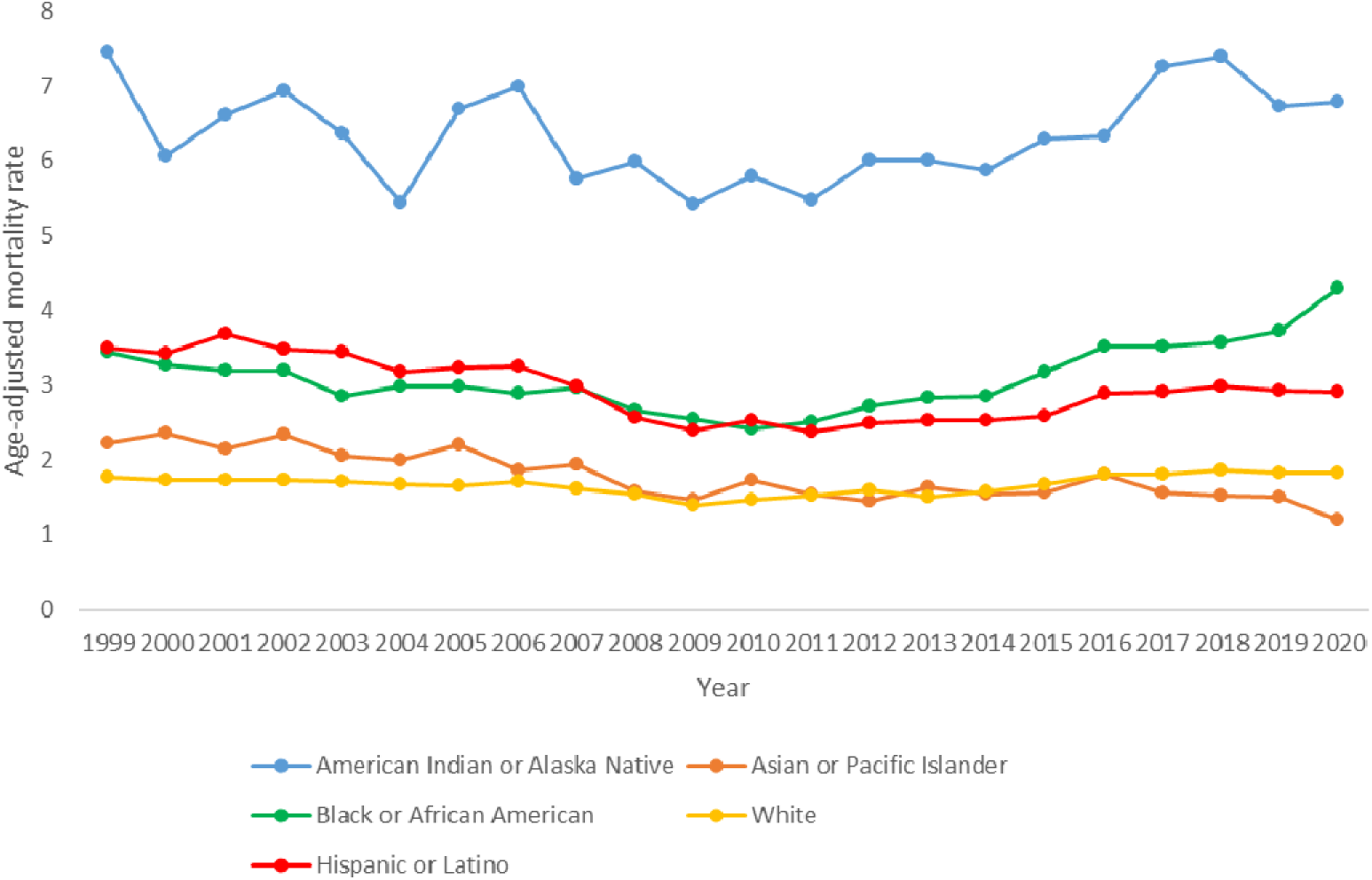
Pedestrian age-adjusted mortality rate stratified by race/ethnicities in the United States; 1999-2020

NH-Black or African American had an AAMR of 3.44 in 1999 and 4.30 in 2020. A declining trend was observed from 1999 to 2011 (APC −2.48; 95% CI −3.24 to −1.72) and a subsequent surge till 2020 (APC 5.75; 95% CI 4.71 to 6.80). **(Table A1, Figure 2, Table A5)**

Hispanics or Latinos had an AAMR of 3.50 in 1999 and 2.90 in 2020. A steady decline was observed from 1999 to 2006 (APC −1.46; 95% CI −3.20 to 0.31) followed by a notable decline from 2006 to 2009 (APC −9.95; 95% CI −20.76 to 2.33) and a steady increase from 2009 to 2020 (APC 2.22; 95% CI 1.46 to 2.99). **(Table A1, Figure 2, Table A5)**

NH-Asian or Pacific Islander had an AAMR of 2.23 in 1999 and 1.19 in 2020. A general declining trend was observed from 1999 to 2020 (APC −2.35; 95% CI −3.01 to −1.68). **(Table A1,** **Figure** 2, Table A5)

NH-White had an AAMR of 1.77 in 1999 and 1.82 in 2020. A downward trend in mortality was observed from 1999 to 2006 (APC −0.57; 95% CI −1.74 to 0.62) which was then followed by a faster rate of decline from 2006 to 2009 (APC −5.17; 95% CI −13.87 to 4.42) followed by an upward trend from 2009 to 2020 (APC 2.57; 95% CI 1.94 to 3.21). **(Table A1, Figure 2, Table A5)**

### Pedestrian-related Crude mortality rate (CMR) stratified by 10-year age groups

Individuals aged *>85 years* had the highest CMR of 5.97 in 1999 and 3.14 in 2020. CMR declined sharply between 1999 and 2009 (APC −3.92; 95% CI −5.33 to −2.51), then steadily between 2009 and 2020 (APC −0.21; 95% CI −1.46 to −1.06). **(Table A1, Figure 3, Table A6)**

**Figure 3:**
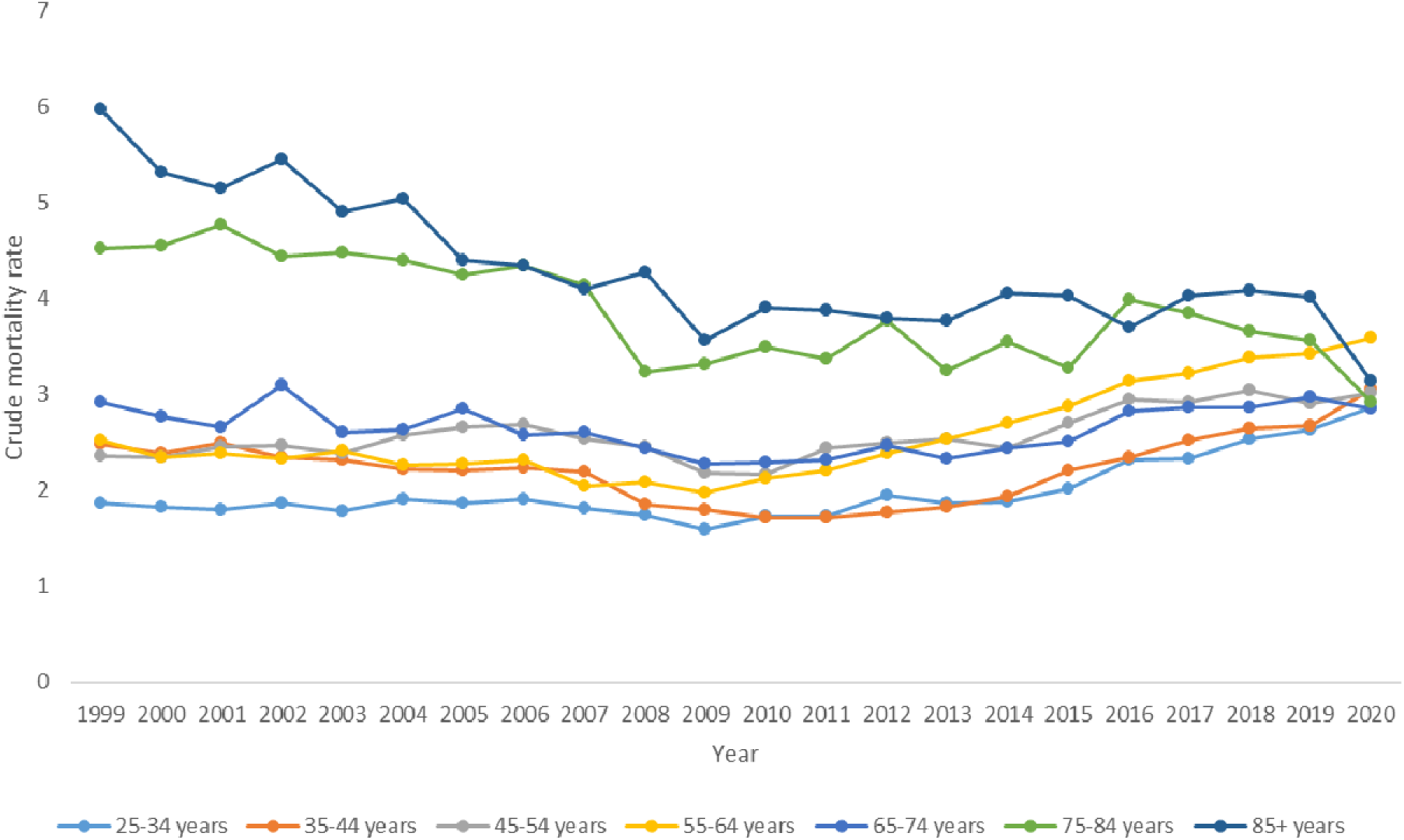
Pedestrian crude-mortality rate stratified by 10-year age-groups in the United States; 1999-2020

Individuals aged *75-84 years* had the second highest CMR of 4.52 in 1999 and 2.92 in 2020. From 1999 to 2013, the CMR decreased (APC −2.68; 95% CI −3.73 to −1.62), then increased from 2013 to 2017 (APC 4.58; 95% CI −6.84 to 17.39), and then declined sharply from 2017 to 2020 (APC −8.09; 95% CI −17.88 to 2.85). **(Table A1, Figure 3, Table A6)**

Individuals aged *65-74 years* had a CMR of 2.92 in 1999 and 2.85 in 2020. A lowering trend in CMR was observed from 1999 to 2011 (APC −1.99; 95% CI −2.97 to −1.02), whereas an increase trend was observed from 2011 to 2020 (APC 2.91; 95% CI 1.60 to 4.24). **(Table A1, Figure 3, Table A6)**

The CMR for people *55 to 64* years was 2.52 in 1999 and 3.59 in 2020. An APC value of −2.24 (95% CI −2.91 to −1.57) corresponded to a downward trend in CMR from 1999 to 2009, and an APC value of 6.18 (95% CI 5.16 to 7.20) resulted in sharp increase in CMR from 2009 to 2017. From 2017 to 2020, there was a consistent rise (APC 3.24; 95% CI 0.34 to 6.22). **(Table A1, Figure 3, Table A6)**

For people aged *45 to 54 years*, the CMR was 2.36 in 1999 and 3.02 in 2020. From 1999 to 2006, CMR showed an increasing trend (APC 2.19; 95% CI 0.46 to 3.94). From 2006 to 2009, there was a sharp decline (APC −6.05; 95% CI −16.62 to 5.86), and from 2009 to 2020, CMR showed an increase in trend (APC 3.11; 95% CI 2.29 to 3.94). **(Table A1, Figure 3, Table A6)**

Individuals aged 35 to 44 years had a CMR of 2.48 in 1999 and 3.06 in 2020. CMR declined from 1999 to 2006 (APC −1.72; 95% CI −2.89 to −0.54), and further declined from 2006 to 2011 (APC −5.65; 95% CI −8.61 to −2.58). From 2011 to 2020 the CMR showed a sharp incline (APC 6.92; 95% CI 5.94 to 7.91). **(Table A1, Figure 3, Table A6)**

The CMR for people in the *25–34 age* group was 1.86 in 1999 compared to 2.86 in 2020. An increase in CMR was observed from 1999 to 2006 (APC 0.63; 95% CI −1.06 to 2.35), then from 2006 to 2009, it decreased (APC −5.76; 95% CI −17.43 to 7.57). Between 2009 and 2020, there was a noticeable increase in the CMR value (APC 5.22; 95% CI 4.46 to 5.98). **(Table A1, Figure 3, Table A6)**

### Pedestrian-related AAMR stratified by Geographic Region

There were discernible differences between the AAMR in the various states. In New Hampshire, the AAMR was 0.99; in New Mexico, it was 3.76. When comparing states in the lower 10th percentile (New Hampshire, Iowa, Vermont, Nebraska, Minnesota, Maine) to those in the top 90th percentile (Mississippi, Louisiana, South Carolina, Arizona, Florida, and New Mexico), the Age-Adjusted Mortality Rate (AAMR) in the higher percentile states was approximately three times higher. **(Table A7, Figure 4)**

**Figure 4:**
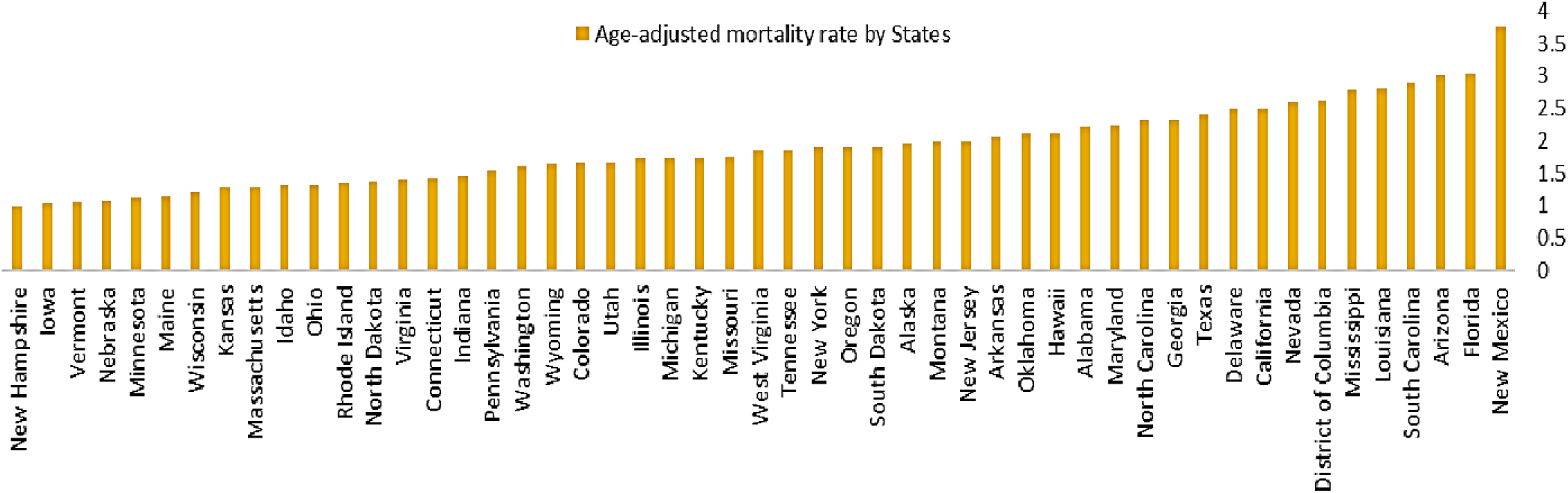
Pedestrian age-adjusted mortality rate stratified by states; 1999-2020

The urban-rural classification based on the 2013 Urbanisation showed that the overall AAMR trends from 1999 to 2020 were similar for both rural areas (86.25) and Medium/Small metropolitan areas (86.67). In contrast, Large Metropolitan’s AAMR was 90.87. Between 1999 and 2006, the large metropolitan showed a negative trend (APC −0.82; 95% CI −1.75 to −0.11), which was followed by a steep decline from 2006 to 2009 (APC −6.39; 95% CI −13.02 to 0.76), and then a rebound in AAMR until 2020 (APC 2.77; 95% CI 2.27 to 3.27). **(Table A1, Figure 5, Table A8)**

**Figure 5:**
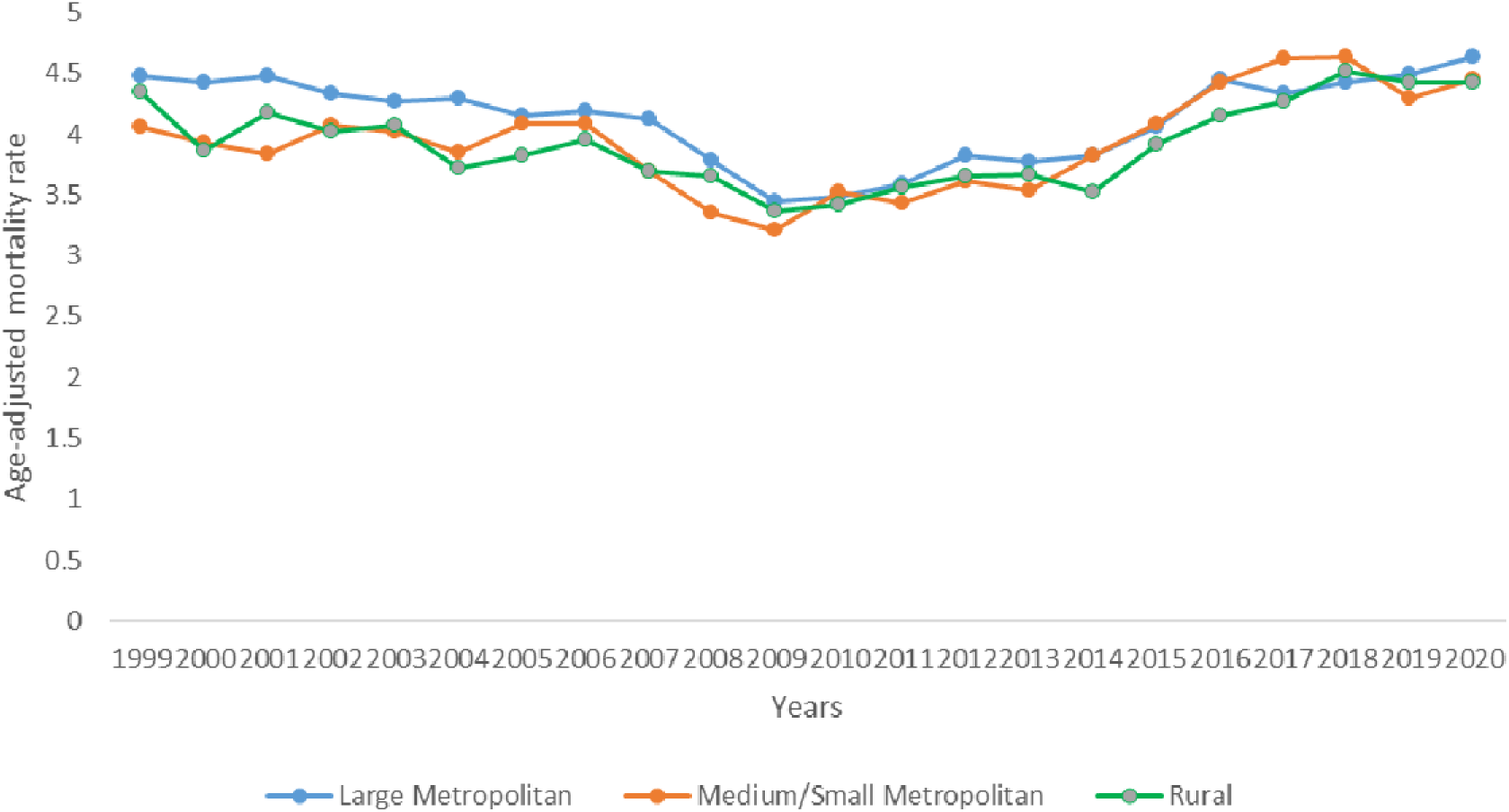
Pedestrian age-adjusted mortality rate stratified by 2013 Urbanization in the United States; 1999-2020.

The mortality trend in medium/small metropolitan areas increased from 1999 to 2006 (APC 0.23; 95% CI −1.35 to 1.83), then declined from 2006 to 2009 (APC −7.68; 95% CI −18.47 to 4.53). From 2009 to 2017, there was a considerable rise in trend (APC 4.58; 95% CI 2.98 to 6.22). Then there was a drop between 2017 and 2020 (APC −0.93; 95% CI −5.93 to 4.34). **(Table A1, Figure 5, Table A8)**

From 1999 to 2011, the Age-Adjusted Mortality Rate (AAMR) in rural regions declined with an APC of −1.65 (95% CI: −2.34 to −0.96). This trend was followed by an increase in AAMR up to 2020, with an APC of 3.23 (95% CI: 2.12 to 4.36). **(Table A1, Figure 5, Table A8)**

## DISCUSSION

To the best of our knowledge, this study is the first to document pedestrian mortality trends in the United States spanning over two decades (1999-2020). A total of 140,280 pedestrian deaths were reported using the CDC-WONDER database. Initially, there was a gradual decrease in the overall AAMR from 1999 to 2006, which was followed by a progressive decline from 2006 to 2009. From 2009 to 2020, there have been a significant increase in the overall AAMR. Males had consistently greater AAMR as compared to females during the course of the study and demonstrated a progressive increase in mortality since 2009. Among races and ethnicities, NH-American Indian or Alaska native had the highest AAMR while NH-White recorded the lowest AAMR. NH-Black or African American reported to have the highest APC (5.75) from 2011-2020. Older adults especially those over 85 years had the greatest crude mortality rates, while highest APC (6.92) was reported in people aged 35-44 from 2011-2020. Significant disparities also exist across the region with states in the top 90^th^ percentile (Mississippi, Louisiana, South Carolina, Arizona, Florida, New Mexico) have almost triple AAMR, than states in the lower 10^th^ percentile. Additionally, large metropolitan areas exhibited greater AAMR than medium/small metropolitan and rural locations. Furthermore, 53.72% of pedestrian deaths occurred in a medical facility (outpatient and inpatient), while a staggering 42.42% deaths occurred in “other” unclassified locations, underscoring the crucial role that healthcare plays in these mortalities.

Our results indicate significant disparities in the AAMR among males and females. with males consistently experiencing higher mortality rates than females. This difference is attributed to a combination of social and behavioral factors that elevate the risk. Evidence indicates that males are more likely than females to engage in unsafe pedestrian behaviours, such as jaywalking and running red lights, which significantly increases their susceptibility to injury and death. [16,17] Additionally, elevated alcohol consumption among males, impairs cognitive function and reaction time further compounding the risk of injury. [18] Furthermore, males tend to focus more on moving vehicles and pay less attention to traffic lights and other pedestrians. This lack of attention to additional traffic cues increases their risk of fatalities at intersections, where comprehensive awareness of the environment is crucial for pedestrian safety. [19] To reduce pedestrian mortality, counter measures holistically targeting pedestrians as well as drivers is crucial. Interventions such as public education campaigns for drivers, augmented visual and auditory cues for pedestrians at crossing signals, and the incorporation of automated pedestrian detection systems are critical. Moreover, infrastructure improvements like raised medians, curb extensions, and enhanced crosswalk markings can further enhance pedestrian safety and mitigate gender-based mortality differences. [20]

Significant disparities in pedestrian mortality are evident across racial and ethnic groups, with NH-American Indians and Alaska Natives being the most vulnerable of all racial/ethnic groups, followed by NH-Black or African Americans. Among NH American Indians and Alaska Natives, higher rates of alcohol consumption compared to other racial and ethnic groups have been identified as a key contributor to increased pedestrian fatalities. [1] Additional alcohol outlets within a neighbourhood are also associated with a 14.2% increase in pedestrian injury risk, suggesting strict regulation of the density of alcohol outlets can be effective in reducing pedestrian injuries [21] Socioeconomic factors further exacerbate this risk: communities with higher poverty rates and lower incomes exhibit elevated pedestrian fatality rates, largely due to increased reliance on walking as a primary mode of transportation and greater exposure to traffic hazards. [22] Additionally, rural areas with higher populations of NH American Indians or Alaska Natives often lack adequate pedestrian infrastructure and enforcement measures, leading to higher mortality rates. [23]

Similar trends are observed among NH Black populations, who disproportionately depend on walking for transportation and are often exposed to unsafe pedestrian environments. [24] Systemic factors, including racial bias in healthcare delivery and disparities in drivers’ yielding behaviors at crosswalks, further contribute to increased pedestrian mortality among marginalized groups. [25,26,27] Annually, approximately 47,000 hospital admissions are attributed to pedestrian-related injuries, disproportionately affecting Black, Hispanic, and multiracial populations. These groups experience higher hospitalization rates, greater healthcare costs per capita, and longer hospital stays, reflecting underlying inequities in transportation safety and healthcare access. [27] Addressing these disparities requires multi-layered strategies that prioritize the social determinants of health. Interventions should include strengthening pedestrian infrastructure in underserved areas, implementing measures to reduce alcohol-related injuries (such as frequent sobriety checkpoints), combating systemic racism in healthcare and traffic systems, and addressing broader socioeconomic inequalities. [28] Targeted, equity-focused approaches are essential to alleviate risk and improve pedestrian safety outcomes among vulnerable populations.

Similarly, elderly individuals experience the highest rates of pedestrian mortality compared to younger age groups. Age-related declines in physical capabilities, slower reaction times, and increased frailty contribute to greater risk of severe injury and death among older pedestrians. [29,30] In contrast, the 35-44 age group has also shown significant increase in APC (6.92) from 2011-2020, possibly reflecting greater exposure to urban traffic environments. Distracted walking from usage of cell phones is possibly linked to impaired situational awareness, increasing crossing duration and risk of injury. [31] Given the increased vulnerability of older pedestrians, who often face longer reaction times and limited assistance navigating traffic; innovative interventions are essential. Potential strategies include adaptive traffic signal systems that extend crossing times, wearable technologies that alert drivers to the presence of elderly pedestrians, and community-based programs that pair seniors with walking companions. [32] In addition, emerging technologies such as advanced pedestrian detection systems and autonomous vehicle innovations are promising in further enhancing pedestrian safety. Rigorous evaluation of these interventions will be critical to informing policies aimed at reducing pedestrian mortality among older adults. [32]

Our results also indicate that Large metropolitan had a higher AAMR than medium/small metropolitans and rural areas. According to the National Highway Traffic Safety Administration, 18% of pedestrian fatalities in 2017 occurred at intersections regardless of crosswalk markings, and approximately 73% of fatalities happened outside designated crosswalks. [33] Moreover, increased nighttime activity in cities further exacerbates pedestrian risks. Research from the AAA Foundation for Traffic Safety indicates that about 30% of serious or fatal vehicle crashes occur between midnight and 3:00 AM, with nighttime fatal crashes being four times more frequent than those during the day [34]. Behavioral factors also significantly contribute to pedestrian injuries. From 2012 to 2014, approximately 18% of road crashes were attributed to negligent or careless driving, often fueled by alcohol impairment or distracted driving, leading to noncompliance with traffic signs and signals [35].

Compounding these risks; the widespread use of larger vehicles such as SUVs in metropolitan areas has significantly worsened pedestrian safety outcomes. SUVs, with their higher front ends, raise the risk of pedestrian fatalities by 22%, especially with impacts at critical injury points like the chest or head [36]. Their larger size also creates substantial blind spots, further diminishing drivers’ ability to detect pedestrians. [36] Addressing these multifactorial risks requires comprehensive urban planning strategies, public safety education, and vehicle design improvements to ensure safer environments for all road users.

Our findings also reveal substantial disparities in pedestrian mortality rates across states, with higher AAMR observed in Florida and New Mexico (98^th^ and 100^th^ percentile) compared to states in lower 10^th^ percentile; Nebraska or New Hampshire. States in the top 90th percentile typically feature large, diverse populations, significant tourist activity, and greater economic disparities, all contributing to increased pedestrian activity and higher injury risk [37]. Large urban centers naturally foster more vehicle-pedestrian interactions, elevating the likelihood of collisions. Interestingly, although rural counties have lower pedestrian activity, individuals injured there are almost four times more likely to die compared to those in urban counties, as shown by a California Department of Public Health analysis of highway patrol reports dating back to 1961 [38]. This elevated fatality rate in rural areas is largely attributed to higher vehicle speeds, delayed emergency response times, limited trauma care access, and poorer infrastructure. Moreover, states with large metropolitan areas and diverse populations face additional risks, as immigrants and tourists unfamiliar with local traffic regulations may engage in unsafe pedestrian behaviors [39]. High population density further amplifies these risks; for instance, Florida; home to four of the five most dangerous cities for pedestrians, has a pedestrian mortality rate (3.9 per 100,000) significantly higher than the national average (2.3 per 100,000) [40]. A study analyzed 37 years of traffic crash data from Florida, attributed the devastating impact of day-light savings on fatal traffic accidents, consequently resulting from disruptions in circadian rhythm of the drivers. [41] Development of rapid bus transit system in Central Avenue, Albuquerque, New Mexico, has significantly decreased (57.1%) risk of fatal injury and collision. [42] These findings suggest; rapid development of Bus Rapid Transit systems across states can prove to be extremely beneficial in making roads and cross walks safer and resourceful for the most vulnerable – the pedestrians.

### Study Limitations

Our study has several limitations. The use of ICD codes for categorizing pedestrian mortality may introduce potential misclassification errors on death certificates Also, the CDC WONDER database lacks specific details on each pedestrian fatality, such as the precise spot of the occurrence, the actions of both the pedestrian and the motorist, traffic conditions, and weather, all of which can have a substantial influence on mortality outcomes. Additionally, the database lacks data on socio-economic factors, pedestrian health status, and, most importantly, access to medical care, which are crucial for understanding disparities in pedestrian mortality rates. Moreover, the data does not include information on the use of safety measures, such as pedestrian crosswalks, traffic lights, and pedestrian visibility aids, which are important factors in pedestrian safety. Also, periodic patterns in pedestrian fatalities may be influenced by changes in reporting techniques or the adoption of innovative security measures that are not included in the data collection. The database also lacks information on emergency response times and the quality of post-collision medical care, both of which are important factors in determining the outcome of pedestrian injuries. When extracting for race/ethnicities, 1059 deaths resulted in the ‘not stated’ section. Hence they were not reported in the results. CDC-WONDER does not provide age-adjusted mortality rates, rather it provides crude rates, when data is stratified by age-groups or place of death. Since this study is a retrospective analysis, the sample size for this study was pre-determined at the time of abstracting pedestrian mortality data from the CDC-WONDER database, therefore the calculation and justification of the sample size cannot be established.

## CONCLUSION

Our comprehensive analysis revealed substantial trends and disparities in pedestrian mortality in the United States from 1999 to 2020. The overall pedestrian AAMR has been increasing since 2009-2020. By focusing on the social determinants of health, particularly systemic racism and access to safe transportation, educational campaigns for both pedestrians and drivers, improvements in infrastructure and targeted efforts to mitigate socio-economic inequalities, we can create more equitable and effective interventions that enhance pedestrian safety and overall community health.

## DECLARATIONS

### 1. Ethics approval and consent to participate

This study was exempt from local institutional review board approval because it uses de-identified government-issued public use data set and follows the STROBE (Strengthening the Reporting of Observational Studies in Epidemiology) guidelines for reporting. “CDC-WONDER is a public service developed and operated by the Centers for Disease Control and Prevention, an agency of United States federal government. The public web site at http://wonder.cdc.gov is in the public domain, and only provides access to public use data and information. You may access the information freely, and use, copy, distribute or publish this information without additional or explicit permission.”

### 2. Availability of data and materials

All data generated or analysed during this study are included in this published article and the Appendix file.

### 3. Competing interests

The authors declare that they have no competing interests.

### 4. Funding

The authors have no funding to disclose for the research, authorship or publication of this article.

### 5. Author Contributions

The author contributions to the paper as follows: *SUA:* Conceptualization, Data curation, Formal Analysis, Validation, Visualization, Writing-review and editing, Supervision. *AK:* Validation, Visualization, Writing-original draft and review and editing. *AT:* Validation, Visualization, Writing-original draft. *MI, AHS*, *SMUD and AI*: Writing-original draft. *AM:* Validation and visualization. All authors have commented on the previous versions of the manuscript and have read and approved the final manuscript.

## Supporting information

Table A1, Table A2 etc.

## Data Availability

All data produced in the present work are contained in the manuscript and in the supplemental files.

http://www.wonder.cdc.gov

## Acknowledgements

All authors would like to sincerely thank the Research Council of Pakistan (RCOP) for their guidance and mentorship

## Notes

### Competing Interest Statement

The authors have declared no competing interest.

### Funding Statement

The study did not receive any funding

### Author Declarations

Ethics committee/IRB approval was waived off because it uses de-identified government-issued public use data set and follows the STROBE (Strengthening the Reporting of Observational Studies in Epidemiology) guidelines for reporting. CDC-WONDER is a public service developed and operated by the Centers for Disease Control and Prevention, an agency of United States federal government. The public web site at http://wonder.cdc.gov is in the public domain, and only provides access to public use data and information. You may access the information freely, and use, copy, distribute or publish this information without additional or explicit permission. All necessary patient/participant consent has been obtained. any patient/participant/sample identifiers cannot be used to identify individuals

