## Supplementary material for "Trends in Pedestrian-related mortality in the United States; 1999-2020, A CDC-WONDER analysis": Table A1, Table A2 etc.

| **Table A1 Annual Percent Change (APC) of Pedestrian age-adjusted mortality rate and crude-**  **mortality rate per 100,000 population in the United States; 1999-2020.** | | | | | |
| --- | --- | --- | --- | --- | --- |
|  |  |  | **APC** | **Lower CI** | **Upper CI** |
| **Overall** |  |  |  |  |  |
| 1999 -2006 |  |  | -0.84 | -1.96 | 0.28 |
| 2006-2009 |  |  | -6.38 | -13.56 | 1.38 |
| 2009-2020 |  |  | 3.10 | 2.60 | 3.61 |
| **Demographics** |  |  |  |  |  |
| *Gender* |  |  |  |  |  |
| **Male** |  |  |  |  |  |
| 1999-2006 |  |  | -0.82 | -1.57 | -0.07 |
| 2006-2009 |  |  | -6.33 | -11.03 | -1.38 |
| 2009-2013 |  |  | 2.04 | -0.68 | 4.83 |
| 2013-2016 |  |  | 5.36 | 0.27 | 10.72 |
| 2016-2020 |  |  | 1.23 | -0.30 | 2.78 |
| **Female** |  |  |  |  |  |
| 1999-2006 |  |  | -0.84 | -1.96 | 0.29 |
| 2006-2009 |  |  | -5.28 | -13.62 | 3.86 |
| 2009-2014 |  |  | 2.20 | -0.88 | 5.37 |
| 2014-2017 |  |  | 5.93 | -2.60 | 15.21 |
| 2017-2020 |  |  | -0.16 | -3.90 | 3.72 |
| **Race and ethnicity** |  |  |  |  |  |
| **NH-American Indian or Alaska Native** | |  |  |  |  |
| 1999-2011 |  |  | -1.65 | -3.02 | -0.26 |
| 2011-2020 |  |  | 2.79 | 0.80 | 4.82 |
| **NH-Asian or Pacific Islander** | |  |  |  |  |
| 1999-2020 |  |  | -2.35 | -3.01 | -1.68 |
| **NH-Black or African American** | |  |  |  |  |
| 1999-2011 |  |  | -2.48 | -3.24 | -1.72 |
| 2011-2020 |  |  | 5.75 | 4.71 | 6.80 |
| **NH-White** |  |  |  |  |  |
| 1999-2006 |  |  | -0.57 | -1.74 | 0.62 |
| 2006-2009 |  |  | -5.17 | -13.87 | 4.42 |
| 2009-2020 |  |  | 2.57 | 1.94 | 3.21 |
| **Hispanic or Latino** |  |  |  |  |  |
| 1999-2006 |  |  | -1.46 | -3.20 | 0.31 |
| 2006-2009 |  |  | -9.95 | -20.76 | 2.33 |
| 2009-2020 |  |  | 2.22 | 1.46 | 2.99 |
| **Age-groups^a^** |  |  |  |  |  |
| 25-34 | 1999-2006 |  | 0.63 | -1.06 | 2.35 |
|  | 2006-2009 |  | -5.76 | -17.43 | 7.57 |
|  | 2009-2020 |  | 5.22 | 4.46 | 5.98 |
| 35-44 | 1999-2006 |  | -1.72 | -2.89 | -0.54 |
|  | 2006-2011 |  | -5.65 | -8.61 | -2.58 |
|  | 2011-2020 |  | 6.92 | 5.94 | 7.91 |
| 45-54 | 1999-2006 |  | 2.19 | 0.46 | 3.94 |
|  | 2006-2009 |  | -6.05 | -16.62 | 5.86 |
|  | 2009-2020 |  | 3.11 | 2.29 | 3.94 |
| 55-64 | 1999-2009 |  | -2.24 | -2.91 | -1.57 |
|  | 2009-2017 |  | 6.18 | 5.16 | 7.20 |
|  | 2017-2020 |  | 3.24 | 0.34 | 6.22 |
| 65-74 | 1999-2011 |  | -1.99 | -2.97 | -1.02 |
|  | 2011-2020 |  | 2.91 | 1.60 | 4.24 |
| 75-84 | 1999-2013 |  | -2.68 | -3.73 | -1.62 |
|  | 2013-2017 |  | 4.58 | -6.84 | 17.39 |
|  | 2017-2020 |  | -8.09 | -17.88 | 2.85 |
| 85+ | 1999-2009 |  | -3.92 | -5.33 | -2.51 |
|  | 2009-2020 |  | -0.21 | -1.46 | -1.06 |
| **2013 Urbanization** |  |  |  |  |  |
| Large Metropolitan | 1999-2006 |  | -0.82 | -1.75 | 0.11 |
|  | 2006-2009 |  | -6.38 | -13.02 | 0.76 |
|  | 2009-2020 |  | 2.77 | 2.27 | 3.27 |
| Medium/Small Metro | 1999-2006 |  | 0.23 | -1.35 | 1.83 |
|  | 2006-2009 |  | -7.68 | -18.47 | 4.53 |
|  | 2009-2017 |  | 4.58 | 2.98 | 6.22 |
|  | 2017-2020 |  | -0.93 | -5.93 | 4.34 |
| Rural | 1999-2011 |  | -1.65 | -2.34 | -0.96 |
|  | 2011-2020 |  | 3.23 | 2.12 | 4.36 |

**a=** indicates APC of crude-mortality rates

**NH =** Non-Hispanic

**Table A2: Absolute number of pedestrian deaths stratified by gender, race/ethnicity in the United States from 1999-2020.**

| **Year** | **Overall** | **Females** | **Males** | **NH - American Indian or Alaska Native** | **NH - Asian or Pacific Islander** | **NH - Black or African American** | **NH - White** | **Hispanic or Latino** |
| --- | --- | --- | --- | --- | --- | --- | --- | --- |
| 1999 | 6108 | 1796 | 4312 | 149 | 182 | 1128 | 3624 | 951 |
| 2000 | 5938 | 1779 | 4159 | 127 | 193 | 1061 | 3525 | 954 |
| 2001 | 6129 | 1801 | 4328 | 148 | 190 | 1087 | 3564 | 1076 |
| 2002 | 6135 | 1868 | 4267 | 149 | 227 | 1083 | 3579 | 1041 |
| 2003 | 6049 | 1809 | 4240 | 149 | 203 | 979 | 3555 | 1104 |
| 2004 | 6028 | 1789 | 4239 | 128 | 218 | 1038 | 3511 | 1078 |
| 2005 | 6117 | 1783 | 4334 | 158 | 244 | 1067 | 3465 | 1146 |
| 2006 | 6208 | 1836 | 4372 | 166 | 216 | 1043 | 3562 | 1185 |
| 2007 | 6003 | 1790 | 4213 | 140 | 230 | 1081 | 3383 | 1127 |
| 2008 | 5628 | 1697 | 3931 | 150 | 201 | 994 | 3249 | 1000 |
| 2009 | 5262 | 1576 | 3686 | 135 | 195 | 971 | 2934 | 980 |
| 2010 | 5500 | 1630 | 3870 | 146 | 243 | 935 | 3106 | 1033 |
| 2011 | 5671 | 1688 | 3983 | 138 | 228 | 981 | 3262 | 1026 |
| 2012 | 6072 | 1852 | 4220 | 153 | 220 | 1086 | 3457 | 1117 |
| 2013 | 6017 | 1797 | 4220 | 158 | 268 | 1157 | 3253 | 1135 |
| 2014 | 6310 | 1853 | 4457 | 160 | 264 | 1180 | 3458 | 1195 |
| 2015 | 6743 | 1966 | 4777 | 171 | 275 | 1334 | 3625 | 1285 |
| 2016 | 7403 | 2170 | 5233 | 169 | 344 | 1463 | 3959 | 1429 |
| 2017 | 7502 | 2264 | 5238 | 199 | 304 | 1505 | 3949 | 1485 |
| 2018 | 7745 | 2334 | 5411 | 200 | 312 | 1566 | 4061 | 1566 |
| 2019 | 7749 | 2295 | 5454 | 180 | 319 | 1640 | 4005 | 1573 |
| 2020 | 7963 | 2323 | 5640 | 179 | 260 | 1909 | 3935 | 1638 |
| Total | 140280 | 41696 | 98584 | 3452 | 5336 | 26288 | 78021 | 26124 |

**Table A3: Pedestrian mortality stratified by place of death in the United States; 1999-2020.**

| **Place of Death** | **Deaths** | **% of Total Deaths** |
| --- | --- | --- |
| Medical Facility - Inpatient | 33323 | 23.75% |
| Medical Facility - Outpatient or ER | 36951 | 26.34% |
| Medical Facility - Dead on Arrival | 4992 | 3.56% |
| Medical Facility - Status unknown | 97 | 0.07% |
| Decedent's home | 2815 | 2.01% |
| Hospice facility | 708 | 0.50% |
| Nursing home/long term care | 1304 | 0.93% |
| Other | 59508 | 42.42% |
| Place of death unknown | 582 | 0.41% |

**Table A4: Pedestrian overall and gender stratified age adjusted mortality rate per 100,000 in the United States from 1999-2020.**

| **Year** | **Overall** | **Female** | **Male** |
| --- | --- | --- | --- |
| 1999 | 2.21 | 1.22 | 3.27 |
| 2000 | 2.10 | 1.21 | 3.10 |
| 2001 | 2.16 | 1.23 | 3.16 |
| 2002 | 2.14 | 1.24 | 3.09 |
| 2003 | 2.07 | 1.20 | 3.05 |
| 2004 | 2.04 | 1.16 | 3.00 |
| 2005 | 2.07 | 1.14 | 3.04 |
| 2006 | 2.05 | 1.17 | 3.03 |
| 2007 | 1.96 | 1.11 | 2.88 |
| 2008 | 1.80 | 1.07 | 2.64 |
| 2009 | 1.69 | 0.97 | 2.44 |
| 2010 | 1.74 | 1.00 | 2.53 |
| 2011 | 1.76 | 1.03 | 2.55 |
| 2012 | 1.88 | 1.12 | 2.72 |
| 2013 | 1.84 | 1.07 | 2.67 |
| 2014 | 1.90 | 1.08 | 2.79 |
| 2015 | 2.02 | 1.14 | 2.93 |
| 2016 | 2.20 | 1.26 | 3.19 |
| 2017 | 2.20 | 1.30 | 3.17 |
| 2018 | 2.25 | 1.32 | 3.23 |
| 2019 | 2.24 | 1.29 | 3.21 |
| 2020 | 2.32 | 1.31 | 3.34 |
| Total | 44.64 | 25.64 | 65.03 |

**Table A5: Pedestrian age adjusted mortality rate per 100,000 stratified by race/ethnicity in the United States from 1999-2020.**

| **Year** | **NH - American Indian or Alaska Native** | **NH - Asian or Pacific Islander** | **NH - Black or African American** | **NH - White** | **Hispanic or Latino** |
| --- | --- | --- | --- | --- | --- |
| 1999 | 7.45 | 2.23 | 3.44 | 1.77 | 3.50 |
| 2000 | 6.06 | 2.35 | 3.26 | 1.73 | 3.43 |
| 2001 | 6.62 | 2.14 | 3.19 | 1.73 | 3.68 |
| 2002 | 6.93 | 2.34 | 3.20 | 1.73 | 3.47 |
| 2003 | 6.36 | 2.05 | 2.86 | 1.71 | 3.45 |
| 2004 | 5.44 | 2.00 | 2.99 | 1.68 | 3.17 |
| 2005 | 6.70 | 2.20 | 2.99 | 1.66 | 3.23 |
| 2006 | 6.99 | 1.87 | 2.88 | 1.71 | 3.25 |
| 2007 | 5.75 | 1.94 | 2.97 | 1.61 | 2.98 |
| 2008 | 5.98 | 1.58 | 2.67 | 1.54 | 2.57 |
| 2009 | 5.42 | 1.47 | 2.54 | 1.39 | 2.39 |
| 2010 | 5.80 | 1.73 | 2.42 | 1.47 | 2.53 |
| 2011 | 5.47 | 1.54 | 2.51 | 1.52 | 2.38 |
| 2012 | 6.01 | 1.45 | 2.71 | 1.60 | 2.49 |
| 2013 | 6.00 | 1.64 | 2.83 | 1.50 | 2.52 |
| 2014 | 5.87 | 1.54 | 2.85 | 1.57 | 2.53 |
| 2015 | 6.30 | 1.55 | 3.17 | 1.67 | 2.59 |
| 2016 | 6.33 | 1.81 | 3.51 | 1.80 | 2.88 |
| 2017 | 7.27 | 1.56 | 3.51 | 1.80 | 2.91 |
| 2018 | 7.40 | 1.52 | 3.57 | 1.87 | 2.99 |
| 2019 | 6.73 | 1.51 | 3.73 | 1.82 | 2.93 |
| 2020 | 6.78 | 1.19 | 4.30 | 1.82 | 2.90 |
| Total | 139.66 | 39.21 | 68.10 | 36.70 | 64.77 |

**Table A6: Pedestrian crude mortality rate per 100,000 stratified by 10-year age groups in the United States from 1999-2020.**

| **Year** | **Age 25-34 years** | **Age 35-44 years** | **Age 45-54 years** | **Age 55-64 years** | **Age 65-74 years** | **Age 75-84 years** | **Age 85+ years** |
| --- | --- | --- | --- | --- | --- | --- | --- |
| 1999 | 1.86 | 2.48 | 2.36 | 2.52 | 2.92 | 4.52 | 5.97 |
| 2000 | 1.82 | 2.39 | 2.34 | 2.34 | 2.77 | 4.54 | 5.31 |
| 2001 | 1.79 | 2.49 | 2.45 | 2.39 | 2.66 | 4.76 | 5.15 |
| 2002 | 1.86 | 2.35 | 2.46 | 2.33 | 3.09 | 4.44 | 5.45 |
| 2003 | 1.78 | 2.31 | 2.39 | 2.41 | 2.60 | 4.48 | 4.90 |
| 2004 | 1.90 | 2.22 | 2.58 | 2.26 | 2.63 | 4.40 | 5.04 |
| 2005 | 1.86 | 2.20 | 2.66 | 2.27 | 2.85 | 4.24 | 4.39 |
| 2006 | 1.90 | 2.23 | 2.69 | 2.31 | 2.58 | 4.34 | 4.34 |
| 2007 | 1.81 | 2.19 | 2.54 | 2.04 | 2.60 | 4.14 | 4.09 |
| 2008 | 1.74 | 1.85 | 2.45 | 2.08 | 2.44 | 3.23 | 4.27 |
| 2009 | 1.59 | 1.80 | 2.18 | 1.98 | 2.27 | 3.32 | 3.56 |
| 2010 | 1.73 | 1.72 | 2.17 | 2.13 | 2.29 | 3.49 | 3.91 |
| 2011 | 1.73 | 1.71 | 2.44 | 2.21 | 2.31 | 3.37 | 3.87 |
| 2012 | 1.94 | 1.77 | 2.49 | 2.38 | 2.47 | 3.77 | 3.79 |
| 2013 | 1.87 | 1.82 | 2.53 | 2.54 | 2.33 | 3.24 | 3.77 |
| 2014 | 1.88 | 1.93 | 2.44 | 2.70 | 2.44 | 3.55 | 4.06 |
| 2015 | 2.02 | 2.20 | 2.70 | 2.88 | 2.51 | 3.28 | 4.02 |
| 2016 | 2.32 | 2.35 | 2.95 | 3.14 | 2.82 | 3.98 | 3.70 |
| 2017 | 2.33 | 2.52 | 2.92 | 3.22 | 2.86 | 3.85 | 4.02 |
| 2018 | 2.54 | 2.64 | 3.04 | 3.39 | 2.87 | 3.66 | 4.08 |
| 2019 | 2.63 | 2.67 | 2.91 | 3.43 | 2.97 | 3.56 | 4.01 |
| 2020 | 2.86 | 3.06 | 3.02 | 3.59 | 2.85 | 2.92 | 3.14 |
| Total | 43.76 | 48.90 | 56.71 | 56.54 | 58.13 | 85.08 | 94.84 |

**Table A7: State-level pedestrian age-adjusted mortality rates per 100,000 people in the United States, 1999-2020.**

| **State** | **Rank** | **Percentile** | **Age Adjusted Rate** |
| --- | --- | --- | --- |
| New Hampshire | 1 | 0 | 0.99 |
| Iowa | 2 | 2 | 1.04 |
| Vermont | 3 | 4 | 1.05 |
| Nebraska | 4 | 6 | 1.07 |
| Minnesota | 5 | 8 | 1.12 |
| Maine | 6 | 10 | 1.15 |
| Wisconsin | 7 | 12 | 1.21 |
| Kansas | 8 | 14 | 1.29 |
| Massachusetts | 8 | 14 | 1.29 |
| Idaho | 10 | 18 | 1.30 |
| Ohio | 10 | 18 | 1.30 |
| Rhode Island | 12 | 22 | 1.35 |
| North Dakota | 13 | 24 | 1.38 |
| Virginia | 14 | 26 | 1.40 |
| Connecticut | 15 | 28 | 1.41 |
| Indiana | 16 | 30 | 1.46 |
| Pennsylvania | 17 | 32 | 1.55 |
| Washington | 18 | 34 | 1.60 |
| Wyoming | 19 | 36 | 1.65 |
| Colorado | 20 | 38 | 1.66 |
| Utah | 21 | 40 | 1.67 |
| Illinois | 22 | 42 | 1.73 |
| Michigan | 22 | 42 | 1.73 |
| Kentucky | 24 | 46 | 1.74 |
| Missouri | 25 | 48 | 1.76 |
| West Virginia | 26 | 50 | 1.85 |
| Tennessee | 27 | 52 | 1.86 |
| New York | 28 | 54 | 1.89 |
| Oregon | 29 | 56 | 1.90 |
| South Dakota | 29 | 56 | 1.90 |
| Alaska | 31 | 60 | 1.97 |
| Montana | 32 | 62 | 1.98 |
| New Jersey | 33 | 64 | 1.99 |
| Arkansas | 34 | 66 | 2.07 |
| Oklahoma | 35 | 68 | 2.10 |
| Hawaii | 36 | 70 | 2.12 |
| Alabama | 37 | 72 | 2.22 |
| Maryland | 38 | 74 | 2.23 |
| North Carolina | 39 | 76 | 2.32 |
| Georgia | 40 | 78 | 2.33 |
| Texas | 41 | 80 | 2.40 |
| Delaware | 42 | 82 | 2.48 |
| California | 43 | 84 | 2.49 |
| Nevada | 44 | 86 | 2.60 |
| District of Columbia | 45 | 88 | 2.61 |
| Mississippi | 46 | 90 | 2.78 |
| Louisiana | 47 | 92 | 2.80 |
| South Carolina | 48 | 94 | 2.88 |
| Arizona | 49 | 96 | 3.02 |
| Florida | 50 | 98 | 3.04 |
| New Mexico | 51 | 100 | 3.76 |

**Table A8: Pedestrian age-adjusted mortality rate per 100,000 mortality stratified by urban-rural classification in the United States; 1999-2020.**

| **Year** | **Large Metropolitan** | **Medium/Small Metropolitan** | **Rural** |
| --- | --- | --- | --- |
| 1999 | 4.48 | 4.06 | 4.35 |
| 2000 | 4.43 | 3.93 | 3.86 |
| 2001 | 4.48 | 3.84 | 4.18 |
| 2002 | 4.33 | 4.07 | 4.02 |
| 2003 | 4.27 | 4.02 | 4.07 |
| 2004 | 4.30 | 3.85 | 3.72 |
| 2005 | 4.15 | 4.09 | 3.83 |
| 2006 | 4.19 | 4.09 | 3.96 |
| 2007 | 4.12 | 3.70 | 3.69 |
| 2008 | 3.78 | 3.36 | 3.66 |
| 2009 | 3.44 | 3.21 | 3.37 |
| 2010 | 3.48 | 3.52 | 3.42 |
| 2011 | 3.59 | 3.43 | 3.56 |
| 2012 | 3.83 | 3.61 | 3.66 |
| 2013 | 3.77 | 3.54 | 3.67 |
| 2014 | 3.83 | 3.83 | 3.53 |
| 2015 | 4.06 | 4.09 | 3.91 |
| 2016 | 4.45 | 4.42 | 4.15 |
| 2017 | 4.33 | 4.62 | 4.27 |
| 2018 | 4.43 | 4.64 | 4.52 |
| 2019 | 4.49 | 4.30 | 4.42 |
| 2020 | 4.64 | 4.45 | 4.43 |
| Total | 90.87 | 86.67 | 86.25 |
